# Mapping the quality of Norwegian health information –Does it facilitate informed choices?

**DOI:** 10.1101/2025.09.09.25335428

**Authors:** Jürgen Kasper, Betül Cokluk, Marianne Molin, Anke Steckelberg, Sandro Zacher, the MAPPinfo project group, Victoria T. Hjellset

## Abstract

**Background and aim:** Health literacy is the ability to use relevant information to make informed choices. However, the quality of the information available will determine how well a person can make those choices. Evidence-based recommendations for the development and design of health information have recently been published. Our study mapped the quality of Norwegian web-based health information in selected public health domains.

**Methods:** Using a multiple-cross-sectional design we assessed information in 16 health domains relevant to infants, children, and youth. Convenience samples were drawn using structured Google searches. Quality appraisal was carried out by three independent raters applying the 19 criteria of the MAPPinfo-checklist. Inter-rater reliabilities were calculated using T-coefficients. Information quality was statistically described. To explain variance of quality, mean quality scores were compared across three independent variables: the type of the health problem, the target group and the provider classes to explain variance of quality.

**Results:** Throughout the surveys in 64 subdomains 1948 health information materials were assessed. Inter-rater reliability was excellent (mean T=.89 /.90). On average, the materials complied with 22% (range 0-73%, SD=.09) of the current minimal standard. Differences between types of problems or target groups were marginal. No differences were found between information provided by health authorities, health services or commercial entities.

**Conclusion:** Norwegian web-based health information is not of sufficient quality to facilitate informed health choices made by citizens. These findings apply to a wide range of public health domains relating to infants, children, and youth. In the absence of appropriate health information of acceptable quality estimates on the public’s levels of health literacy might need to be reconsidered. Further research is needed to appraise the quality of information in other health domains and countries.

## Background

The age of evidence-based practice has redefined the responsibility for making health-related decisions. Previously, specialists such as doctors, but also health authorities, were expected to identify and recommend the right course of action among several possible alternatives. However, these days ordinary citizens – the lay public-are encouraged and allowed to make their own health decisions, depending on the type of the decision either in cooperation with health care professionals or independently. This can only work well in terms of good health outcomes if good quality information is made available to them, so that they are well-informed about their choices [1,2]. Informed choices are based on relevant knowledge, consistent with the decision-maker’s values and behaviourally implemented [2].

Reflecting a consensus within scientific communities and many societies worldwide, the “informed choice” has become an important quality parameter of healthcare provisions, and is anchored in international ethical standards [3,4], national patient rights [5], and many health professional guidelines [6]. This implies that health service users, such as patients in a hospital, visitors of municipal health care centres, or national health information platforms, expect to receive help in making informed decisions about their health. This help includes forms of communication that encourage and support the understanding and intellectual processing of information relevant to the pending decision [7,8].

Many, or even most, health-related decisions are made by people themselves without involving healthcare providers and, therefore, potentially without them even becoming formal health service users. Examples are choices made about whether or not to visit the doctor about a health concern, using a bicycle or a car to get to work, or to using or not using hormonal contraception. Most people seek health-related information through Internet search engines and do not base their decisions solely upon information provided by health professionals [9].

The way, people search, select, and use information generally varies between individuals, and is guided by intuitive rather than rational processes. The former are shaped by emotional states, social contexts, economic conditions, motivations, preferences, literacy and various intellectual abilities [10]. The unique nature of these intuitive processes provides opportunities for companies such as Google or Meta to systematically manipulate decision-making behaviours by analysing individual search patterns and then influencing their choices by changing their personal search algorithms.

The quality of health choices is also influenced by individual competencies to evaluate the quality and reliability of the information, and avoid being influenced by outside forces, in order to make choices that will lead to good health outcomes – subsumed under the term health literacy [11]. A recent review showed that health literacy was found to be low in the Norwegian population society [12]. As a result of these findings, the Norwegian government has developed a strategy to strengthen health literacy in its population [13]. Amongst other anticipated benefits of these efforts, Norwegian people might make better use of existing health information and establish healthier lifestyles. However, for this to occur, at least some information sources of high quality must be available.

Information that facilitates the making of informed choices is called evidence-based health information (EBHI) [14]. “Evidence-based” refers to both the content and presentation of information. Decades have been spent on researching how to develop and design health information in order to facilitate informed health choices. This evidence is summarized in the guideline “Evidence-based health information” [15] which provides evidence updates on 21 research questions regarding the development and design of health information. In addition, the guidelines summarize the previously published ethical guidelines [4]. Specifically, the guideline’s recommendations relate to transparency with regard to the background of the authors and the origin of the content, the completeness of the content, its presentation considering knowledge about sources of bias or misunderstanding, the methods of information search, selection and evaluation used during development, and, finally, measures to involve the target group in developing the guideline, and documentation of evaluation of the suitability of the information for the target group [15].

Having in mind the current emphasis on health literacy and its definition as a competency to process information, we believe, that the most exciting question to ask is whether citizens of our country even do have suitable information available they could process. Our project focuses on information about health problems which do not necessarily require healthcare provider involvement to be managed and are relevant for the target group of public health nurses (PHN). Norwegian public health nursing is a unique service addressing children and youth (aged 0 – 20 years) and their families with a health-promoting and preventive approach. Working on a municipal basis in schools and health care centres, PHNs play a crucial role in education and empowerment of the children, youth and families, so that they make well-informed health decisions for themselves. With regard to multiple health issues relevant for these target groups, the function of PHN implies to be accountable for counselling and information provision if needed, however, not necessarily having a formal role in making the related health decision.

Robust evidence already exists around the evaluation of health information provision using various concepts of quality and evaluation methods in general [16–23]. In a previous study, we searched for evaluation methods complying with evidence-based quality criteria but could not find any [24]. We therefore assume, that no systematic evaluation of health information quality has been conducted based on the current guidelines [15]. Moreover, we are certain that this type of research has not been conducted in Norway where we are affiliated.

This study aimed to map the quality of Norwegian web-based health information materials (WBHIMs) in selected public health domains relevant to the clientele of Norwegian public health nurses. Specifically, it aimed to determine whether Norwegian people can make informed health choices based on the information available on the Internet. The study also aimed to provide insight into the nature of any potential shortcomings. Moreover, three independent variables – type of the health problem, provider class and target group – were tested for a potential contribution to explaining variance regarding the quality of Norwegian health information.

## Methods

### Design

The study used a multi cross-sectional design mapping the quality of on the Internet openly accessible health information in selected health domains relevant for the clientele of Norwegian public health nursing services. The term ‘health domain’ is used to label superordinate topics, such as sleeping problems of infants or perimenstrual complaints. Regarding the research design, the scrutiny was focused on the level of sub-domains, such as prevention of sudden infant death, treatment of delayed onset of sleep, or treatment of menstrual pain or extensive bleeding. Each subdomain was defined in analogy with a medical indication by, firstly, a specific diagnosis, secondly, the type of measures (diagnostic, treatment, prevention, health promotion or rehabilitation), and, thirdly, a particular target group. Each single cross-sectional study analysed the quality of all identifiable WBHIMs in a specific subdomain.

The current analysis integrates the findings of 64 cross-sectional studies in 16 health domains (Table 1), which were part of the MAPPinfo project and carried out by 21 master’s students in two separate cohorts one year apart, using the same method. Focussing on the reality of a user in need of information about a particular health problem, this study was designed to take the user’s perspective. Taking this perspective informed the decisions we made about which health domains and subdomains to focus on. We also developed the search strategy used to identify the websites with regard to how we assumed a user would proceed. Finally, taking the user’s perspective is reflected in the choice of the concept of quality [15] implying informed health choices to be the crucial target [2].

**Table 1:**
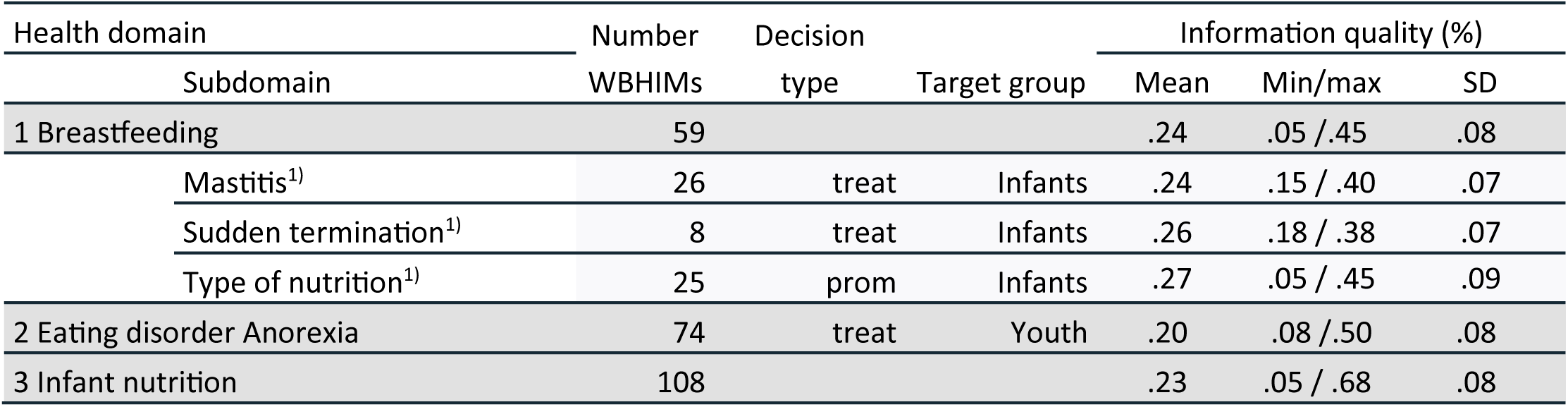
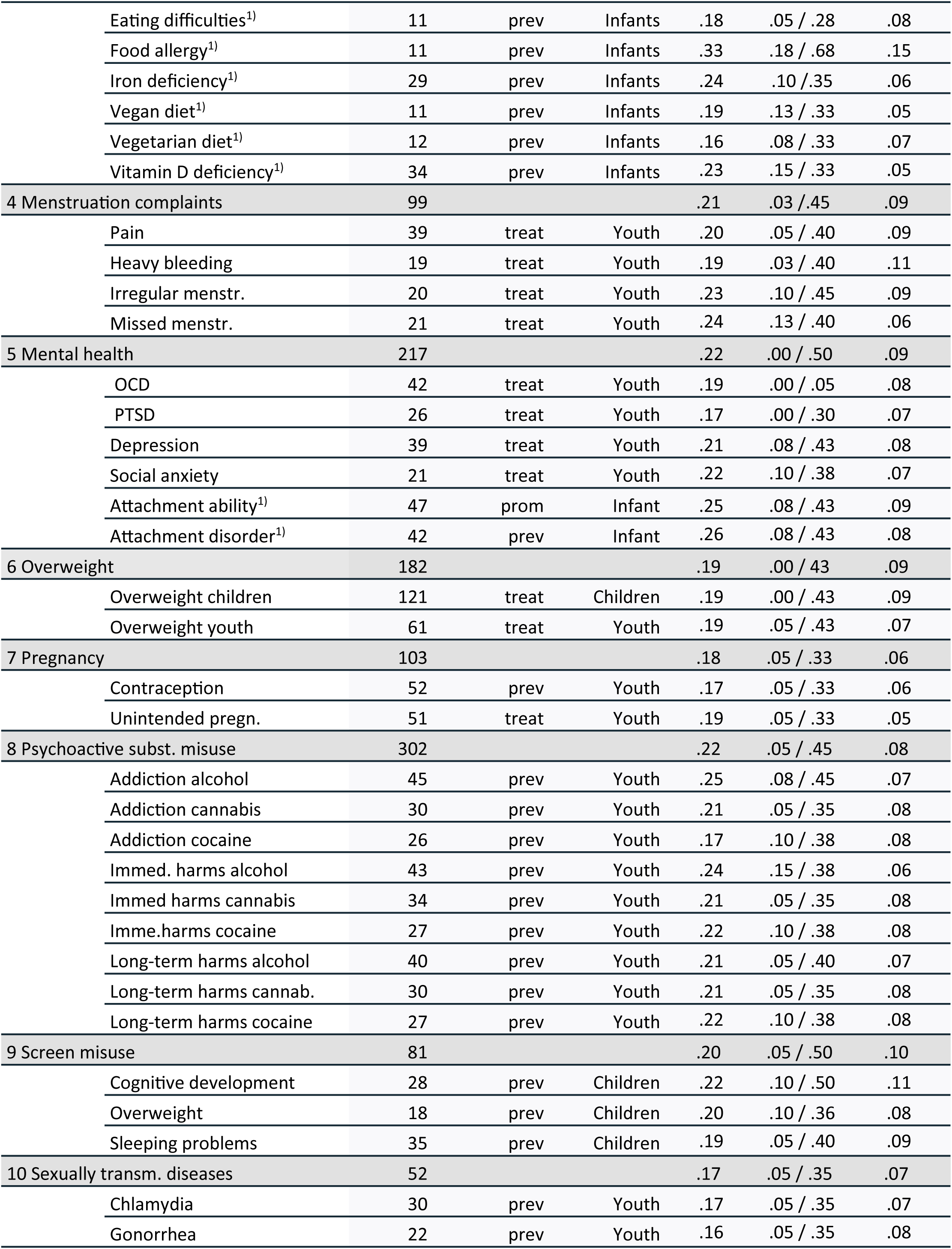
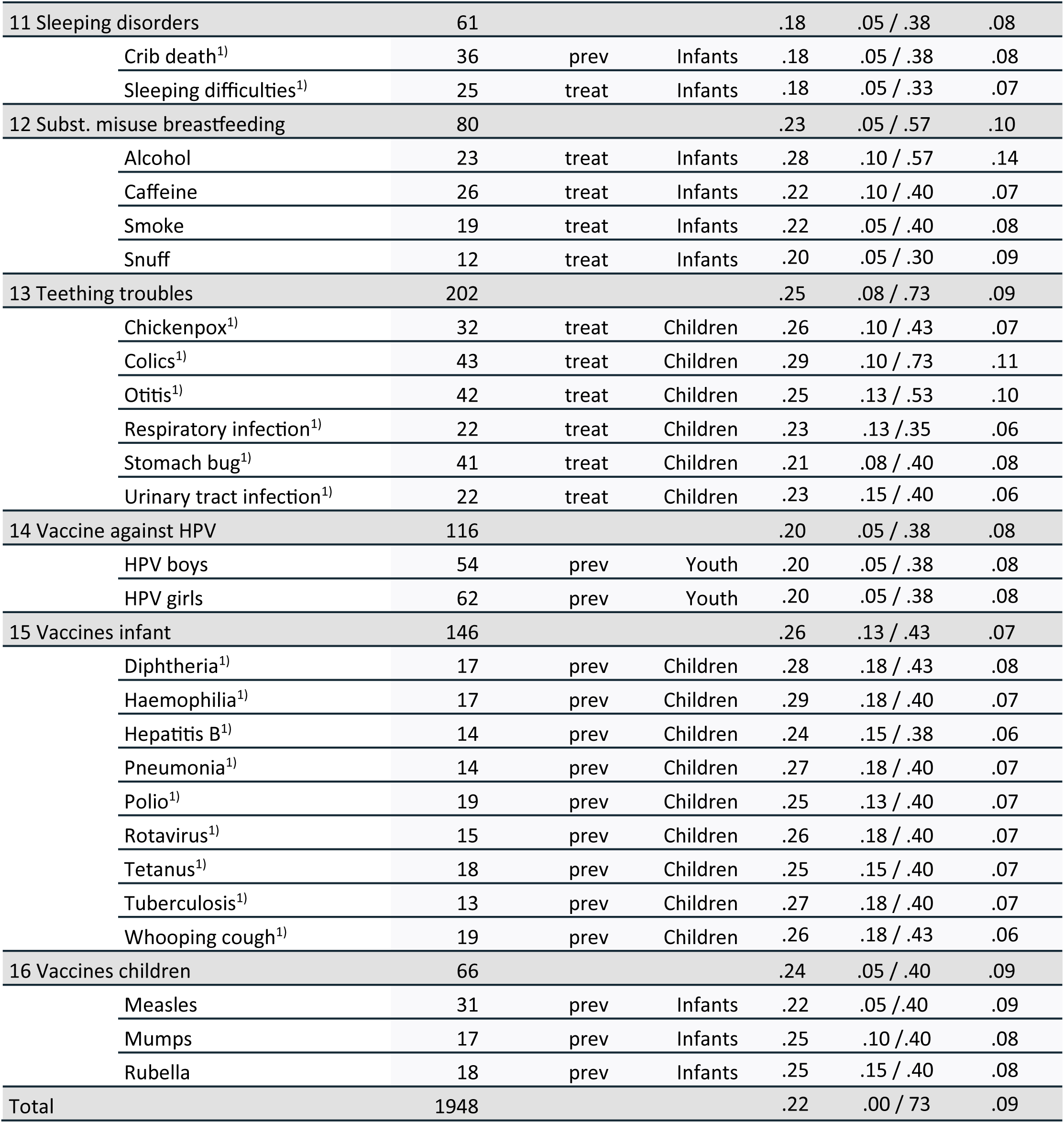
Description of the sample and results on information quality. Table 1: Legend of Table 1: description: prev = information about a problem related to prevention of a specific health state or outcome, treat= information about a problem related to treatment of a specific health state or disease, prom= information about how to promote a specific health state or outcome, WBHIM= web-based health information material. 1)=survey was conducted by the first cohort of master‘s students.

| Health domain | Subdomain | Number WBHIMs | Decision type | Target group | Information quality (%) |  |  |
| --- | --- | --- | --- | --- | --- | --- | --- |
|  |  |  |  |  | Mean | Min/max | SD |
| 1 Breastfeeding |  | 59 |  |  | .24 | .05 / .45 | .08 |
|  | Mastitis<sup>1)</sup> | 26 | treat | Infants | .24 | .15 / .40 | .07 |
|  | Sudden termination<sup>1)</sup> | 8 | treat | Infants | .26 | .18 / .38 | .07 |
|  | Type of nutrition<sup>1)</sup> | 25 | prom | Infants | .27 | .05 / .45 | .09 |
| 2 Eating disorder Anorexia |  | 74 | treat | Youth | .20 | .08 / .50 | .08 |
| 3 Infant nutrition |  | 108 |  |  | .23 | .05 / .68 | .08 |

Table 1: Legend of Table 1: description: prev = information about a problem related to prevention of a specific health state or outcome, treat= information about a problem related to treatment of a specific health state or disease, prom= information about how to promote a specific health state or outcome, WBHIM= web-based health information material.
|  |  |  |  |  |  |  |
| --- | --- | --- | --- | --- | --- | --- |
| Eating difficulties<sup>1)</sup> | 11 | prev | Infants | .18 | .05 / .28 | .08 |
| Food allergy<sup>1)</sup> | 11 | prev | Infants | .33 | .18 / .68 | .15 |
| Iron deficiency<sup>1)</sup> | 29 | prev | Infants | .24 | .10 / .35 | .06 |
| Vegan diet<sup>1)</sup> | 11 | prev | Infants | .19 | .13 / .33 | .05 |
| Vegetarian diet<sup>1)</sup> | 12 | prev | Infants | .16 | .08 / .33 | .07 |
| Vitamin D deficiency<sup>1)</sup> | 34 | prev | Infants | .23 | .15 / .33 | .05 |
| **4 Menstruation complaints** | **99** |  |  | **.21** | **.03 / .45** | **.09** |
| Pain | 39 | treat | Youth | .20 | .05 / .40 | .09 |
| Heavy bleeding | 19 | treat | Youth | .19 | .03 / .40 | .11 |
| Irregular menstr. | 20 | treat | Youth | .23 | .10 / .45 | .09 |
| Missed menstr. | 21 | treat | Youth | .24 | .13 / .40 | .06 |
| **5 Mental health** | **217** |  |  | **.22** | **.00 / .50** | **.09** |
| OCD | 42 | treat | Youth | .19 | .00 / .05 | .08 |
| PTSD | 26 | treat | Youth | .17 | .00 / .30 | .07 |
| Depression | 39 | treat | Youth | .21 | .08 / .43 | .08 |
| Social anxiety | 21 | treat | Youth | .22 | .10 / .38 | .07 |
| Attachment ability<sup>1)</sup> | 47 | prom | Infant | .25 | .08 / .43 | .09 |
| Attachment disorder<sup>1)</sup> | 42 | prev | Infant | .26 | .08 / .43 | .08 |
| **6 Overweight** | **182** |  |  | **.19** | **.00 / .43** | **.09** |
| Overweight children | 121 | treat | Children | .19 | .00 / .43 | .09 |
| Overweight youth | 61 | treat | Youth | .19 | .05 / .43 | .07 |
| **7 Pregnancy** | **103** |  |  | **.18** | **.05 / .33** | **.06** |
| Contraception | 52 | prev | Youth | .17 | .05 / .33 | .06 |
| Unintended pregn. | 51 | treat | Youth | .19 | .05 / .33 | .05 |
| **8 Psychoactive subst. misuse** | **302** |  |  | **.22** | **.05 / .45** | **.08** |
| Addiction alcohol | 45 | prev | Youth | .25 | .08 / .45 | .07 |
| Addiction cannabis | 30 | prev | Youth | .21 | .05 / .35 | .08 |
| Addiction cocaine | 26 | prev | Youth | .17 | .10 / .38 | .08 |
| Immed. harms alcohol | 43 | prev | Youth | .24 | .15 / .38 | .06 |
| Immed harms cannabis | 34 | prev | Youth | .21 | .05 / .35 | .08 |
| Imme.harms cocaine | 27 | prev | Youth | .22 | .10 / .38 | .08 |
| Long-term harms alcohol | 40 | prev | Youth | .21 | .05 / .40 | .07 |
| Long-term harms cannab. | 30 | prev | Youth | .21 | .05 / .35 | .08 |
| Long-term harms cocaine | 27 | prev | Youth | .22 | .10 / .38 | .08 |
| **9 Screen misuse** | **81** |  |  | **.20** | **.05 / .50** | **.10** |
| Cognitive development | 28 | prev | Children | .22 | .10 / .50 | .11 |
| Overweight | 18 | prev | Children | .20 | .10 / .36 | .08 |
| Sleeping problems | 35 | prev | Children | .19 | .05 / .40 | .09 |
| **10 Sexually transm. diseases** | **52** |  |  | **.17** | **.05 / .35** | **.07** |
| Chlamydia | 30 | prev | Youth | .17 | .05 / .35 | .07 |
| Gonorrhea | 22 | prev | Youth | .16 | .05 / .35 | .08 |
| 11 Sleeping disorders | 61 |  |  | .18 | .05 / .38 | .08 |
| Crib death<sup>1)</sup> | 36 | prev | Infants | .18 | .05 / .38 | .08 |
| Sleeping difficulties<sup>1)</sup> | 25 | treat | Infants | .18 | .05 / .33 | .07 |
| 12 Subst. misuse breastfeeding | 80 |  |  | .23 | .05 / .57 | .10 |
| Alcohol | 23 | treat | Infants | .28 | .10 / .57 | .14 |
| Caffeine | 26 | treat | Infants | .22 | .10 / .40 | .07 |
| Smoke | 19 | treat | Infants | .22 | .05 / .40 | .08 |
| Snuff | 12 | treat | Infants | .20 | .05 / .30 | .09 |
| 13 Teething troubles | 202 |  |  | .25 | .08 / .73 | .09 |
| Chickenpox<sup>1)</sup> | 32 | treat | Children | .26 | .10 / .43 | .07 |
| Colics<sup>1)</sup> | 43 | treat | Children | .29 | .10 / .73 | .11 |
| Otitis<sup>1)</sup> | 42 | treat | Children | .25 | .13 / .53 | .10 |
| Respiratory infection<sup>1)</sup> | 22 | treat | Children | .23 | .13 / .35 | .06 |
| Stomach bug<sup>1)</sup> | 41 | treat | Children | .21 | .08 / .40 | .08 |
| Urinary tract infection<sup>1)</sup> | 22 | treat | Children | .23 | .15 / .40 | .06 |
| 14 Vaccine against HPV | 116 |  |  | .20 | .05 / .38 | .08 |
| HPV boys | 54 | prev | Youth | .20 | .05 / .38 | .08 |
| HPV girls | 62 | prev | Youth | .20 | .05 / .38 | .08 |
| 15 Vaccines infant | 146 |  |  | .26 | .13 / .43 | .07 |
| Diphtheria<sup>1)</sup> | 17 | prev | Children | .28 | .18 / .43 | .08 |
| Haemophilia<sup>1)</sup> | 17 | prev | Children | .29 | .18 / .40 | .07 |
| Hepatitis B<sup>1)</sup> | 14 | prev | Children | .24 | .15 / .38 | .06 |
| Pneumonia<sup>1)</sup> | 14 | prev | Children | .27 | .18 / .40 | .07 |
| Polio<sup>1)</sup> | 19 | prev | Children | .25 | .13 / .40 | .07 |
| Rotavirus<sup>1)</sup> | 15 | prev | Children | .26 | .18 / .40 | .07 |
| Tetanus<sup>1)</sup> | 18 | prev | Children | .25 | .15 / .40 | .07 |
| Tuberculosis<sup>1)</sup> | 13 | prev | Children | .27 | .18 / .40 | .07 |
| Whooping cough<sup>1)</sup> | 19 | prev | Children | .26 | .18 / .43 | .06 |
| 16 Vaccines children | 66 |  |  | .24 | .05 / .40 | .09 |
| Measles | 31 | prev | Infants | .22 | .05 / .40 | .09 |
| Mumps | 17 | prev | Infants | .25 | .10 / .40 | .08 |
| Rubella | 18 | prev | Infants | .25 | .15 / .40 | .08 |
| Total | 1948 |  |  | .22 | .00 / .73 | .09 |

The STROBE guideline [25] was applied in the reporting of the current study.

**Fig 1:**
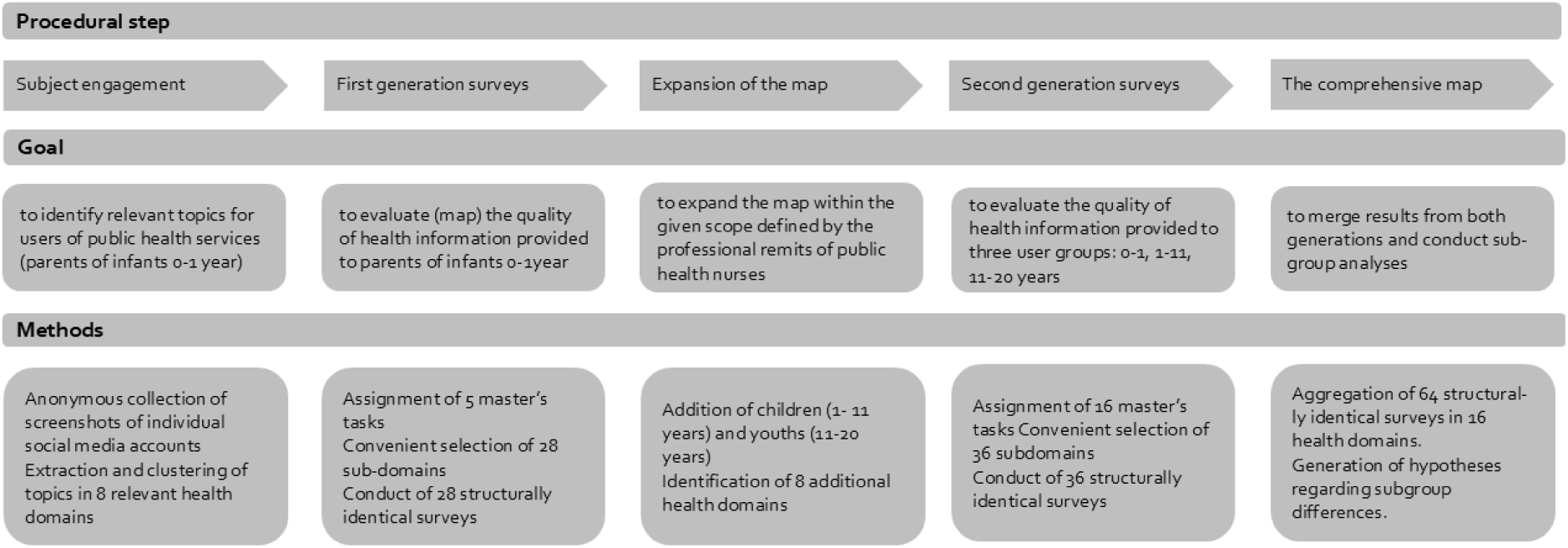
Overview over the procedure of the multi-cross-sectional study. **Fig 1: Legend**: This figure illustrates the procedural steps followed during the conduct of 64 structurally identical surveys.

### Sample

Considering that it would not be possible, necessary, or even meaningful to appraise all WBHIMs existing in the chosen sector of the Norwegian health information landscape, we established a system for the selection of health domains and particular subdomains from the broader map. Choices about which health domains to screen needed to avoid arbitrariness and instead were designed to reflect the users’ experience of relevance.

In the first cohort of 28 surveys conducted by master’s students, we chose domains related to infant health younger than 12 months. To achieve an understanding of which themes might be considered relevant the target group, we conducted a pre-study.

In that pre-study, a convenience sample of twenty parents with children younger than 12 months were contacted via the students’ personal networks and local health care centres. The parents were asked to collect screenshots of health information or health-related claims that they were exposed to through social media over a period of five days (in January 2023). Participants were provided with a study information sheet including instructions and a link to an online form. By sharing their screenshots anonymously online, the parent-participants provided implicit informed consent to use their data in the research. This information was kept confidential, and the authors had no access to any information that could identify individual participants. This pre-study was approved by the Norwegian agency for shared services in education and research (SIKT [26], reference number 768954).

Through this pre-study we collected 228 screenshots, after removing duplicates. The pool of screenshots was analysed by the research group and clustered according to health domains in the given age range. The choice of subdomains to be mapped by the first cohort of studies was inspired by this material, as the students chose from the pool and specified and customized their individual theses. In the second cohort of additional 36 surveys, we extended this scope of the target topics to include health domains related to children between 1 and 11 years of age and youth between 12 and 20 years of age. Identification of relevant domains related to children’s and youth’ health was achieved partly by extrapolating from similar relevant infant domains, such as vaccination or consumption of psychotropic substances, and partly by identifying additional domains with specific relevance to these age ranges, such as pregnancy.

Sampling in the current multi cross-sectional study refers to 64 populations of WBHIMs, according to the number of 64 sub-domains screened. As a common approach across all 64 cross-sectional studies, our recruitment strategy was designed to identify as many as possible WBHIMs that might be hit in a typical Google search by a user.

After clearing the cache of the browser on their computers, the students drew convenience samples of WBHIMs by applying the following selection criteria: Norwegian language, non-professional, and no barriers to access. WBHIMs were identified via structured searches in Google which varied between the 64 single surveys, however, were developed following a common procedure: Starting with search terms likely for a user to choose, we conducted orienting searches with varying combinations. Results were compared using a test set of WBHIMs to learn about redundancy between the terms, and to identify the core terms for the final search. Second, the final search was run. The final strategy also included a cutoff criterion on how many of the references generated by Google search should be considered.

### Measurement methods

Quality appraisal was achieved using the MAPPinfo instrument (Mapping the quality of health information [27]), which is based on the guideline EBPI [15]. The instrument is novel in three regards: 1. It is the first one precisely operationalizing the EBPI guideline recommendations [15,24]. 2. Appraisal is conducted based on the published information material only, without a need to search secondary sources. 3. The instrument can be used reliably by persons without special training or background in evidence-based practice making quality appraisal more transparent and open for everyone.

Following a “pars pro toto” approach, MAPPinfo evaluates only a selection of the criteria provided in the guideline. In particular, MAPPinfo includes criteria which are based on strong recommendations. Also, it does not include criteria which cannot easily be observed in health information documents or websites. Criteria relating to methods of development, or the state of evaluation of a WBHIM, are not included, because they cannot be assessed just based on the in WBHIMs. MAPPinfo has been validated as a valid screening instrument and has been shown to provide a very good estimate of the overall quality of information in reference to the EBPI guideline recommendations [27].

MAPPinfo is designed as a checklist, comprising 19 criteria which are thoroughly described and defined in a manual, which also includes good practice examples. The criteria refer to the following four categories: definitions (how target group, topic, and goal of an information are identified and described), transparency (how accurately and clearly the authors, producers, funding sources, conflicts of interest and origin of information are presented), content (how comprehensive the information provided is in relation to prevalence, natural course, benefits and harms of all alternatives, test security in case of diagnostic options and uncertainty), and presentation (whether the methods used to provide information are evidence-based; particularly methods to communicate quantitative information like effects of treatments).

Although some of the checklist criteria require consideration of several elements, the answering format is just dichotomous or trichotomous giving little space to grade the assessment of quality. This rigorous manner of judging is chosen by the developers of MAPPinfo taking into account the users’ need, to find all elements of a criterion fulfilled to really be able to use the information. If e.g., the benefit is explained complying with the guidelines in terms of absolute risk reduction for only one of three available options, the respective rating would be zero, because information on benefit is considered useless for the citizen until he or she can compare between the options. A grading of quality would potentially better indicate the providers’ skill level; while taking the perspective of a citizen, we need to answer the question, whether or not an informed choice is possible. MAPPinfo represents the essential elements of what is known about health information quality and therefore provides a minimum standard evaluation. There will truly be other potentially relevant criteria, we do not know yet. This is to say, less can never be enough, while full compliance does not necessarily make a good health information.

### Data collection

All websites were classified according to three variables: The type of the health problem was divided into diagnostic –, treatment –, preventive, and topics related to health promotion. In accordance with the sampling structure, three clusters for target groups were differentiated: infants (0 – 12 months), children (1 – 11 years) and youth (12 – 20 years). Health information providers were classified as research-, governmental-, health service-, non-governmental (NGOs), or commercial entities, news organisations, or bloggers and influencers.

Quality appraisal of the WBHIMs was carried out via a rigorous process that ensured consistency in the way the criteria were applied across the 64 separate surveys that were conducted. This was considered appropriate in the context of a research study, despite a previously existing evidence that the checklist’s psychometric properties are sufficient for use by untrained raters [27].

Ratings were documented in a specially prepared MS EXCEL spreadsheet [28], which allowed data to be entered from different raters, including the documentation of areas of disagreements. The spreadsheet generated result tables which contained reliability, information quality, and a diagram which provided a visual representation of the results. Before starting the data collection, the concerned master’s students and the supervising researchers (BC, VTH, JK) met, to calibrate how they were going to apply the criteria consistently to the specific health problem, using examples of the WBHIMs. Beyond strengthening inter-rater reliability this meeting was also supposed to contribute to harmonizing the application of the method over the range of 64 single surveys.

Decisions were also made about which content would be considered as part of a particular WBHIM vs which content would be considered to be external. These decisions were generally made based on the URL of a website. For example, an information website including hierarchically organized pages under a specific landing page and from the same provider would be considered as one WBHIM, whereas a link to a neighbouring website under another landing page or provided by another entity would not be seen as part of that same WBHIM. The WBHIMs were rated consecutively, and independently, by two master’s students, and any disagreements in the ratings were then resolved by discussion. To provide a reference standard, the websites were also rated by an expert in evidence-based health information (BC, VTH, JK), and any disagreements between the students’ consensus and the reference standard were resolved by discussion, before a second consensus rating was documented. The first four of the 64 cross-sectional studies were conducted between February and April 2023 at the Arctic University of Norway in Tromsø at OsloMet University in Oslo, and the final 60 studies were conducted between March and May 2024 at OsloMet.

### Analyses

Data were partly analysed at the level of subdomains using MS EXCEL (version 2402) and then transposed into an SPSS (version 28.0.1.1) file to be analysed on a broader level. Analyses were based on five data columns, one and two comprising the individual ratings by the student raters, column three representing the agreement between the students’, column four comprising the reference standard, and column five, the final consensus.

#### Reliability and validity

To provide an estimate of the quality of the measurement process, inter-rater-agreements were quantified. Regardless of which particular coefficient is chosen, this calculation consists of adjusting the number of observed matches for the number of matches that would occur by chance. Inter-rater reliabilities between the students (column 1 and 2) were calculated on criterion level using the “T” coefficient [29], which is an adjusted Cohen’s kappa [30], however, estimates probabilities of the rate of expected matches not empirically derived from the given data set but theoretically based on the potential number of values a criterion can get. This adjustment of the original Cohen’s kappa reflects the assumption that the rater should orient observations with regard to what is theoretically possible and unbiased by previous observations [31]. Inter-rater-reliability coefficients can score from 0 to 1 and were considered moderate between 0.40 and 0.60, strong if higher than 0.60, and excellent if higher than 0.80 [32]. Means of inter-rater-reliability were calculated for subdomains and rater teams.

The same statistics used when comparing the two student raters as a unit (column 3) and the reference standard (column 4), were also used to provide an estimate of validity of the preliminary results (first consensus). In the absence of a gold standard, the reference standard is considered suitable to calculate a criterion validity. This was done on the level of the single criteria and also as mean validity scores. Due to additional expert appraisal and discourse after the first consensus, these validity scores are considered a very conservative estimation of the validity of the final results (column 5).

#### Quality of the information

Each WBHIM was given a quality score (QS), representing a percentage (0-100) of criteria met. As each single criterion is operationalising one evidence-based strong recommendation, the set of criteria is considered a minimum standard. Since even a violation of one of the criteria can of impede a fully informed choice 100% of the criteria need to be met to facilitate informed choices [27]. In addition, we calculated criteria scores providing the percentage of WBHIMs within a subdomain complying with each specific criterion. The quality scores are reported as means aggregated over all WBHIMs belonging to specific subdomains, domains, and the total sample of WBHIMs. Criteria scores are reported as means aggregating WBHIMs of the total sample.

#### Inference testing of subgroups

In order to provide a better insight into how information quality varies between materials targeting different groups, types of health problems, or being published by different groups of provided, three ANOVAS were performed. All of them used the quality score as the dependent variable. In the first ANOVA, the independent variable was the target group which comprised three factor steps (infants, children, youths), in the second ANOVA the independent variable was the type of health problem which comprised three factor steps (prevention, treatment, health promotion), and in the third ANOVA the independent variable was the provider class which comprised 8 factor steps (research organisations, governmental entities, health service institutions, NGOs, commercial entities, news organs or bloggers/influencers). Effects between within the sets of multiple factor steps were examined more closely using posthoc Scheffé’s tests for pairwise comparisons of factor steps. P-values lower than 0.05 were considered significant. Additional interpretation was guided by the corresponding eta-square parameters estimating the effect size of each p-value. This was particularly important as these inference tests were overpowered and the number of single tests quite high implying the risk of p-value inflation as well as random effects due to multiple testing.

## Results

The complete datafile is accessible at https://zenodo.org/ [33]

### Description of the sample

Through the 64 cross-sectional studies in 16 health domains, we assessed in total 1948 WBHIMs presented on 1538 websites (Table 1). Many websites were evaluated several times with a focus on different subdomains.

Commercial providers represented 45% (876) of the total sample, news and NGOs each represented 13% (256 /259), public health services represented 10% (203), governmental entities represented 9% (184), scientific organisations represented 5% (103) and bloggers represented 3% (67).

### The quality of the measurement

Inter-rater-agreement was excellent in mean for the 64 sub-domains (T=.89, SD=.07) and for 59 of the 64 single subdomains (T min =.73, T max =.99). Criterion validity was excellent in mean for the 64 sub-domains (T=.90, SD =.06, T min=.57, T max =.99) and for 62 of the 64 single subdomains. The sampling of information sites was quite evenly distributed over the target populations (20 infants, 19 children, 25 youth). Subdomains dealing with prevention were strongly represented (37 out of 64 subdomains), treatment related subdomains proportionally (25 out of 64 subdomains) and information on health promotion rarely (2 out of 64 subdomains).

### The quality of the information (descriptive results)

In this chapter we address the first two research questions: 1. What is the level of information quality in the selected health domains with reference to the currently applicable quality criteria defined as a minimal standard? Seen across the 1948 WBHIMs evaluated in the current study, health information is complying on average with 22% of the quality criteria (SD.09, min 0%, max 73%) (Table 1). This implies that WBHIMs are found to disregard 78% of the minimal standards of quality on average. Table 1 provides results in detail for each of the investigated health domains and subdomains.

2. The second research question sought to clarify the nature of information quality within the given samples. This required a focus on the role of each individual quality criterion. Results answering this question are provided in detail in figure 2 and Table 2. The range of percentages by which single criteria are met varies from 0% to 82% over the total sample (Table 2, Fig 2). The three most frequently met criteria are: “target group named” (55%), “language neutral” (72 %), and “use of narratives avoided” (82%). Conversely, three examples of criteria met in fewer than 1% of WBHIMs are: “systematic search strategy reported”, “benefits and harms disclosed”, and “gain loss framing used”.

**Fig 2:**
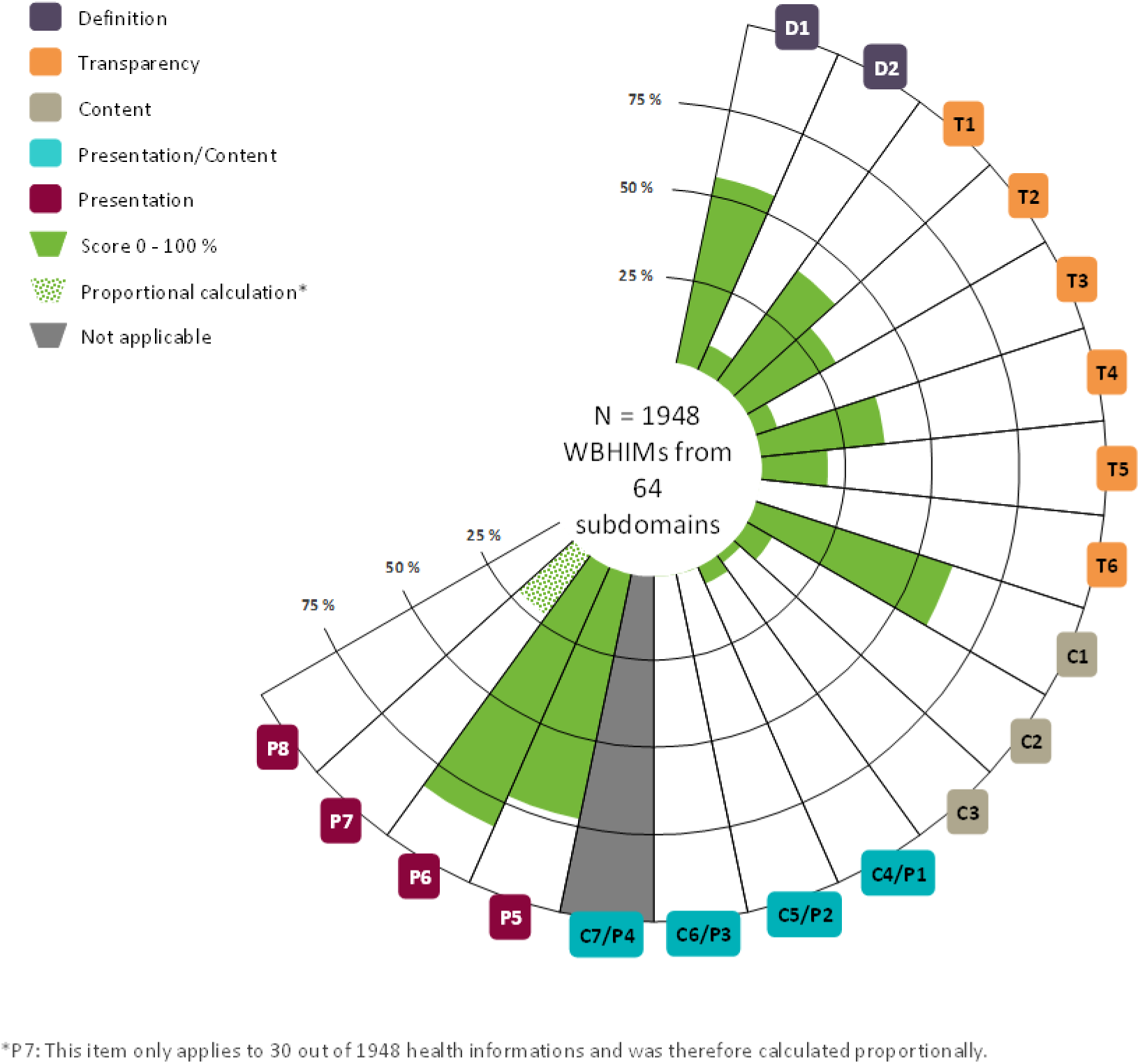
Visualisation of information quality as measured with the MAPPinfo-checklist. **Fig 2: Legend of Fig 2**: The figure visualises the spectrum of quality criteria of the MAPPinfo checklist. The shape of the open circle diagram indicates the nature of the underpinning concept of quality which is not complete (further research will lead to completion of the circumplex model). The green colouring gives a visual analogue impression of the extent of compliance with the respective criteria.

**Table 2:** Information quality according to criteria the 19 MAPPinfo criteria. Table 2: Legend of Table 2: The table provides numbers (percentages and absolute numbers) for the health information materials’ compliance with each of the 19 criteria, which are assigned to categories in the MAPPinfo-checklist. Compliance is quantified as a percentage of the total number of 1948 information items. “n.a.”= not applicable.

| Category |  | Criterion | % met<br>(absolute<br>number) | Std.<br>Deviation |
| --- | --- | --- | --- | --- |
| DEFINITIONS | D1 | Target group of HI | 55(1071) | 50 |
|  | D2 | Aim to facilitate informed choice | 8(156) | 21 |
| TRANSPARENCY | T1 | Author indicated | 40(779) | 44 |
|  | T2 | Funding declared | 30(584) | 46 |
|  | T3 | COI considered | 7(136) | 25 |
|  | T4 | Plan for update | 36(701) | 26 |
|  | T5 | Use of references | 20(390) | 32 |
|  | T6 | Reported syst. search | <1(4) | 4 |
| CONTENT | I1 | Health problem explained | 60(1169) | 49 |
|  | I2 | Options named & explained | 9(171) | 22 |
|  | I3 | Stochastic uncertainty explained | 3(58) | 17 |
| CONTENT /<br>PRESENTATION | I4/P1 | Natural course / prevalence | 6(117) | 23 |
|  | I5/P2 | Benefits presented adequately | <1(11) | 8 |
|  | I6/P3 | Harms presented adequately | <1(10) | 7 |
|  | I7/P4 | Presentation of test quality | n.a. |  |
| PRESENTATION | P5 | Neutral language | 72(1403) | 45 |
|  | P6 | Narratives not used | 82(1597) | 39 |
|  | P7 | Graphics designed suitably | 22(429) | 39 |
|  | P8 | Gain/loss framing | 1(2) | 3 |

### Moderators of information quality (results from inference testing)

This chapter reports results answering the third research question about seeking to explain variation of the quality score within the total sample. As lined out in Table 1, the standard deviation was very low (0.09) indicating that the amount of variation across the 1948 quality scores around the mean value (0.22) is low.

There was a significant main effect for target group, however, associated with a small effect size (p<.001, eta-square=.05). Posthoc pairwise comparisons showed that health information relating to infants (n=745 of 1948) has a slightly higher quality than information relating to children (n=570). Information relating to youth (n=633) has even lower quality compared with each of the two other groups (QS_infant_= 24% min/max: 5/73; QS_children_=21% min/max: 0/50; QS_youth_= 20%, min/max 0/73; all pairwise comparisons are significant: p<.001).

Similarly, there was a significant main effect for decision type, however, associated with a very small effect size (p=.004, eta-square=.006). According to pairwise posthoc Scheffé’s tests, the effect refers to a higher quality of information about problems relating to health promotion (n=72), compared to information relating to treatment (n=875, p=.006) or prevention (n=1001, p=.032) (QS_health promotion_=25% min/max:5/45; QS_prevention_=22%, min/max: 5/68; QS_treatment_=21% min/max: 0/73).

Finally, there was a significant main effect for provider class, however, associated with a small effect size (p<.001, eta-square=.05) (absolute numbers are provided in Table 3). According to pairwise posthoc Scheffé’s tests, information provided by scientific entities have higher quality scores compared to each of the other six provider classes (p<.001). Information from NGOs has higher scores than information from the news (p<.001), however, is not superior to any of the other five provider classes. Governmental information has slightly higher quality scores than information from the news (p=.012), however, is not superior to any of the other 4 provider classes (commercial, health services, news, and bloggers & influencers). There are no differences in quality between information provided by health service entities compared to commercial entities, the news, or bloggers & influencers.

**Table 3:** Information quality by provider classes. Table 3: Legend of Table 3: The table provides numbers indicating the distribution of WBHIMs (=Web-based health information materials) between seven provider classes and quality scores (expressed as %) as absolute mean values for provider classes, standard deviations and the empirical range of quality scores.

| Provider class | N of WBHIMs | QS in % | Std. Deviation | Min/max |
| --- | --- | --- | --- | --- |
| scientific | 103 | 28 | 7 | 13/50 |
| NGO | 256 | 23 | 8 | 05/57 |
| governmental | 184 | 23 | 6 | 05/40 |
| commercial | 876 | 22 | 9 | 00/73 |
| health services | 203 | 21 | 6 | 05/68 |
| news | 259 | 19 | 8 | 03/50 |
| bloggers / influencers | 67 | 19 | 7 | 05/38 |
| Total | 1948 | 22 | 9 | 00/73 |

## Discussion

### Short summary of aim and approach

The current study was designed to investigate whether openly accessible web-based information in the Norwegian language is of good enough quality to enable citizens to make informed choices about their health.

Employing 64 cross-sectional studies we evaluated a total of 1948 WBHIMs in 16 public health domains relevant for young people from birth to legal age. The health problems selected for this study can be managed by the citizen alone without to involve health care providers. The MAPPinfo-checklist, which operationalizes current ethical and evidence-based standards (EBHI [15]), was used as the evaluation method.

The health information investigated in the current study, is on average meeting only 22% of the criteria constituting the minimal standard. Not one of the information material items was close to reaching the minimum standard. Crucial content, such as the prevalence of a health problem that should be prevented, or potential benefits and harms of available treatment alternatives, is regularly missing. These results make us very doubtful that Norwegian citizens are able to find the quality of information they require to make evidence informed health choices.

Information quality varies only slightly between the target groups and types of health problems, and even less between some of the provider classes. Taking into account the very small effect sizes, any differences might have only been statistically significant because of the large size of the sample. Thus, it appears that the very poor quality of information is ubiquitous.

### Limitations

Despite the large number of single surveys combined and the number of websites screened in total, the current study was still able to examine only a small area of the entire information landscape.

Generalisability of these findings therefore needs to be interpreted with caution. Mapping of information quality in other health domains and languages is ongoing and will soon contribute to further completion of the picture.

It could be argued that evaluating information used by the studies’ target groups when making health choices should have included social media. Indeed, social media is now being used for information purposes to an increasing extent [17]. Even governments are starting to rely more on social media to ensure their online information is up to date and accessible [17]. Google as a search engine is, however, still highly relevant in active information seeking. Also, given the general lack of essential meta information in social media, such as, author, funding, source, conflict of interest, publication date, we argue that our results might overestimate, rather than underestimate, the quality of a broader range of sources, including social media.

Selecting WBHIMs via the method we chose (ie. stepwise development and conduct of Google searches followed by the screening of references), may have led to some degree of selection bias of an uncertain nature. However, the risk for bias is likely to be low, because the samples were large and close to completely capturing the respective population of information on that topic. In addition, multiple methodological arrangements were employed to minimize the influence of subjectivity, e.g., the use of the incognito modus in Google, and the consensus discussions within the research group about whether websites should, or should not, be included. The recruitment strategy was designed to mimic the likely search behaviour of non-professional users. So, if we missed any relevant websites, this might have been due to their limited visibility or accessibility.

It could be argued that our evaluation is referring to presentation quality only, as we did not attempt to verify the accuracy of the content, such as numerical information about benefits and harms provided on the websites. By omitting evaluation of its accuracy, good information might have unjustly been devalued due to focusing on presentation weaknesses. This argument implies that it is principally possible to determine whether an information is correct. This position would suggest the existence of an authority that defines the general truth and appears to entrench itself in an outdated paradigm. Evidence-based practice, however, shows that there may be many ways to answer a health question depending on the methods used in the evaluation of the available evidence. Given that it is now generally accepted that there is no such thing as a single form of “truth” about a specific topic information quality is instead defined by the information’s level of transparency and trustworthiness.

The latter is best evaluated by the scientific methods, reasoning, and conclusions involved in the development of the material [27,34,35]. Therefore, MAPPinfo does not guide the evaluator to verify the accuracy of any of the health information content. The checklist is, instead, designed to enable an assessment of the material’s trustworthiness, and a valid judgement of the quality of the information material’s content and presentation [27].

The set of quality criteria employed in this study is not exhaustive of all possible evaluative criteria. In addition to the 19 criteria of MAPPinfo and the guideline EBHI, there is an unknown number of other criteria of potential relevance. However, these criteria are yet to be identified and articulated. The concept of health information quality is still an evolving field of science. New criteria may need to be added, and existing criteria adjusted as the EBHI Guidelines themselves are updated.

It might be regarded as inappropriate to evaluate websites in terms of a particular concept of quality, which providers or developers had not intended to adhere to. It could be further argued, that investigating websites dedicated to facilitating informed choices of their readers, may have led to less dire results. Indeed, our sample did include a high percentage of commercial websites and information from providers not specifically known for prioritizing evidence-based health choices. However, the focus of our study was to explore the quality of the information that Norwegian citizens are exposed to in reality. Thus, we could not select only those sites aiming specifically to provide high quality information. However, none of the sources of information that we evaluated met the minimum quality standards. This applies to information from all kinds of providers, falsifying our hypothesis of a higher quality of information delivered from the health authorities and health care services.

### Results in the context of the literature

First, our results need to be discussed in the context of other research on quality of health information. Insufficient quality of health information has previously been documented in hundreds of studies carried out in multiple countries and languages regardless of the particular medical domain. Most of these studies deal with readability and understandability [36], fewer with credibility and accuracy [37,38], some refer to evidence-based criteria [39,40]. We did not find any studies evaluating health information via the use of the quality criteria recommended in the guideline EBHI [15]. A recent study on officially recognized disease awareness campaigns, such as the “Breast Cancer Awareness Month” or “Movember” (for men’s health) found the respective websites highlighted the benefits more than the harms of any health interventions mentions and rarely discussed potential problems such as overdiagnosis or overtreatment. They tended to use an unbalanced approach, encouraging readers to make certain choices, rather than neutrally informing them about the availability of common tests and treatments [41]. The current study has mapped the quality of a large range of information topics within the field of public health problems, which could be used independently by readers to make their own choices independently of any health care professionals. Further mapping is ongoing within other domains. However, given the ubiquitous nature of our findings, we do not expect to find any substantial differences in the information quality in other health domains.

A look into the particular genre of “Patient decision aids” (PDA,) which is a subset of health information provision, does at least demonstrate that providing high quality information is possible [22,23,42]. PDAs have been frequently evaluated, and some of the results have been impressive [42,23]. However, a rigorous focus on evidence-based criteria has not been applied systematically in all of these studies [24].

The Norwegian Directory of health adopted the International Patient Decision Aid Standard (IPDAS [43]) criteria in 2018 as a quality standard for PDAs published on the Norwegian health platform helsenorge.no [44]. Although not operationalizing the single criteria satisfactorily and using some criteria which are not yet evidence-based [24], the IPDAS overlaps to a large extent with the MAPPinfo checklist. However, no studies have yet been published which evaluate whether Norwegian information materials comply with any of these criteria.

Our results need to be discussed in the context of health literacy. The term is currently receiving increased attention in many countries and health systems [11–13,45–47]. For example, the Norwegian directory of health has made the improvement of its citizens’ health literacy the highest priority [13].

This was in response to comprehensive surveys carried out in Norway and other countries which indicated worryingly low levels of health literacy amongst their citizens [12]. Health literacy is the ability to find, process and use health information [11]. But how can health literacy be adequately assessed in the absence of appropriate information? Our study’s findings suggest that trying to assess health literacy in the absence of a robust quality of health information material is not possible, and the conclusions regarding people’s level of health literacy –as they are published in the afore-mentioned reports-might be based on a misconception. This implies that a national strategy to strengthen the users’ health literacy [13] needs to consider the requirement of appropriate health information too.

Our results on the quality of health information accessed by the lay public should also be discussed in the context of the quality of information used by healthcare providers. Depending on the case and type of the health problem, health care providers are responsible for ensuring that they educate themselves in other ways in order to compensate for the poor quality of health information available on the Internet. This might particularly apply when it comes to specialist medical care, where healthcare providers are directly accountable as decision makers along with the patient.

The types of health problems examined in the current study can be handled independently by the lay public. However, according to their health professional’s guidelines, Norwegian PHN are responsible for the users’ education in the respective health domains [49], suggesting that the PHNs are accountable for compensating for insufficient information quality by supplementing to and correcting information, where the user could not manage alone. When providing health consultations, PHNs are, like other health professionals, strongly encouraged to base their advice on the relevant medical guidelines. But are these guidelines always reliable sources of good quality information? At least as far as the Norwegian guidelines are concerned, this does not seem to be the case. A recent systematic review of Norwegian scientific medical guidelines investigated their compliance with international standards for trustworthy clinical practice [49]. The author group used the 15 criteria of the NEATS instrument [33], which are similar to the MAPPinfo criteria. Most of the NEATS criteria score very low on average across all the Norwegian guidelines. The lowest scores are given to the criteria “study selection”, “description of studies and results”, “grading of effects reported” and “external review”. The highest scores were given for the criterion, “providing unequivocal recommendations”, which in the absence of transparency regarding the justification of such recommendations, might be considered as another barrier to providing good quality information to empower the public to make informed choices.

In summary, the information sources that specialists rely on are not necessarily any more trustworthy than the health information that is made available online to the general public. Consequently, improving the health literacy of HCPs should also be a priority for the Norwegian Directory of Health [48].

### Implications for practice

The low quality of health information available to citizens does not seem to be a superficial problem that could be corrected by asking the information providers to consider a few more of the quality standards. Information developers might need further training in order to understand the published guidance and to implement this knowledge into their development processes. The outcomes of this sort of education process were evaluated in a recent German randomized study [50]. Unfortunately, in this study, such training did not lead to the development of higher quality health information [51].

Considering the evidence on the quality of medical guidelines in Norway [49] a solution to this problem might need to go even deeper. To provide a reliable basis for any health information development, rethinking the procedures and management of evidence updates involved in guideline development seems necessary, at least in the Norwegian context. In an ideal world, the development of medical guidelines for an audience of health professionals should automatically include the development of a partner guideline for the lay public too. For example, as demonstrated by the SHARE-IT project, where patient information is automatically linked to the corresponding medical guideline [52]

As a basic requirement, the agreement on the national information quality standards to be used as a template for health information developments needs to be renewed or updated [44]. There is currently little evidence that the standards determined for Norwegian PDAs are used or even understood by information content developers. The scope of application of the quality standards in charge needs to be expanded to include health information in general, rather than just PDAs.

A related issue is determining how non-governmental providers can be motivated to use the same standards, even though they obviously provide information based on various goals (not all are dedicated to informed choice). A possible solution of this problem might be the implementation of a certification system, giving all types of health information providers the opportunity to learn about, and adhere to, minimal information quality standards via a voluntary code of conduct.

## Conclusions

Regardless of the information provider, Norwegian health information is not of sufficient quality to facilitate the making of informed health choices by Norwegian citizens. This finding is based on the current evidence-based guidelines for health information and applies throughout a wide range of public health domains relevant to infants, children, and youth. More research is needed to investigate the quality of information provided in other health domains and in other countries. The disclosed deficiencies are fundamental and concern all facets of quality, particularly content and presentation.

In the absence of health information of acceptable quality, levels of health literacy in the public might be difficult to assess, and efforts currently being undertaken to strengthen health literacy of Norwegian citizens premature.

## Data Availability

The complete datafile is accessible online at https://zenodo.org/records/16642974

## Acknowledgments

The members of the «MAPPinfo project group» were students (or external supervisors) involved in conducting master theses as part of the project “Quality of health information: the missing link in the era of health literacy”: Helene Kolnes^1^, Hilde Laholt (supervisor) ^1^, Solveig Wergeland Hansen^2^, Petter Christiansen Halse^2^, Frida Kristine Jansen Broen^2^, Amalie Mesel^2^, Lars Peder Kolås Henriksen^2^, Signe Svendsrud Sørlien^2^, Ingrid Thorsen^2^, Hanne Nesse^2^, Christine Lilledahl^2^, Charlotte Myrvollen^2^, Nadia Maria Adablah Teigland^2^, Linn Therese Gunnerød^2^, Marte Næsgaard Myhre^2^, Anna Frydenlund^2^, Synnøve Skattebu^2^, Mari Hårstad^2^, Maiken Jacobsen^2^, Anna Marie Liljenvall^2^, Maren Køhn^2^, Marte Kristiansen^2^.

Affiliations:

1. The arctic University of Tromsø, UiT, Institute of Healthcare and Health Promotion, Faculty of Health Sciences. Tromsø, Norway

2. Oslo Metropolitan University, OsloMet, Institute of Nursing and Health Promotion, Faculty of Health Sciences, Oslo, Norway.

This group is led by the first and corresponding author.

We are very grateful for professional and thoughtful language editing by Dr. Wendy Longley.

## Author contribution statement

Conceptualisation and methodology: Jürgen Kasper, Anke Steckelberg, Victoria Telle Hjellset, Marianne Molin

Recruitment and Data collection: Jürgen Kasper, Sandro Zacher, the MAPPinfo research group, Victoria Telle Hjellset, Betül Cokluk

Analyses: Jürgen Kasper, Sandro Zacher

Writing of the manuscript: Jürgen Kasper, Betül Cokluk

Substantial editing and discussion of the manuscript: Marianne Molin, Anke Steckelberg, Sandro Zacher, the MAPPinfo research group, Victoria Telle Hjellset

All the authors have read and approved the final version of the manuscript.

